# Enhancing fracture detection in wrist radiographs via paired synthetic data generation

**DOI:** 10.1101/2025.09.18.25336124

**Authors:** Stanley A Norris, Daniel Carrion, Sergio Uribe, Mohamed K Badawy

## Abstract

**Purpose:** Cast artefacts in follow-up wrist radiographs reduce diagnostic image quality and complicate the assessment of fracture healing, displacement, and complications. This study evaluated whether targeted image augmentation and cast suppression can improve automated fracture detection performance, particularly when training datasets underrepresent cast images.

**Methods:** A previously published CycleGAN model was repurposed to generate synthetic paired datasets for Pix2Pix training. These Pix2Pix models learned image-to-image translation from synthetic cast to cast-less images. Fracture detection experiments assessed the impact of CycleGAN-based synthetic data augmentation and cast suppression across training sets with varying cast image proportions. Performance was assessed using mean average precision at intersection over union thresholds (mAP@0.5, mAP@0.5:0.95) with bootstrap resampling.

**Results:** Synthetic augmentation with CycleGAN significantly improved fracture detection compared to models trained without any cast images. However, models trained with limited real cast data outperformed those using synthetic augmentation. The Pix2Pix model consistently outperformed CycleGAN-based cast suppression. Cast suppression only improved fracture detection when applied to models that had never been exposed to cast images during training; when applied to models already trained on cast-containing images, suppression decreased accuracy.

**Conclusion:** By generating a synthetic paired dataset, we addressed a critical limitation in cast suppression research and enabled more robust and meaningful architecture comparisons. CycleGAN-based augmentation and cast suppression preprocessing improved fracture detection performance only in models that had not been exposed to casts during training, demonstrating their potential value when real cast data are unavailable. However, models trained with real cast data consistently outperformed those trained with synthetic alternatives, highlighting that real cast data remain essential for developing reliable and clinically robust fracture detection models.

## Introduction

Wrist injuries are a frequent issue across age groups, accounting for approximately 2.5% of emergency department presentations [1]. Radiography is routinely used for diagnosing and evaluating wrist fractures due to its accessibility, affordability, and low radiation exposure [2]. However, damaged limbs often require stabilisation using splints or casts made of resin, fibreglass, or plaster [3]. In follow-up radiographs monitoring the position or healing of fractures, the presence of a cast introduces artefacts that obscure anatomical detail and complicate diagnostic interpretation. This issue is especially pertinent in elderly patients, where the prevalence of osteoporosis increases both fracture risk and the challenge of diagnosis within casts [4, 5]. Frequent monitoring in patients at risk of delayed healing is also hindered by casts, which can impair the radiological assessment of callus formation and signs of non-union, potentially delaying the diagnosis of complications [6, 7].

The clinical need for image quality improvement in follow-up imaging has driven interest in artificial intelligence-based solutions. Few studies have explored cast artefact suppression in extremity radiographs [8–10]. These studies have shown that AI models can be trained to enhance images by suppressing the appearance of cast shadow artefacts in both paediatric and adult wrist x-rays. One study showed improvements in automated fracture detection [9], and another showed improvements in the diagnostic confidence of radiologists reporting on difficult cases [10]. The published studies are subject to limitations across data collection, modelling approaches, and evaluation methods. The use of CycleGAN models introduces risks of anatomical distortion or hallucination that can be difficult to detect or quantify [11]. Performance evaluations based on histogram-derived quantitative metrics such as correlation, intersection, Chi-squared distance, and Hellinger distance are not robust indicators of true diagnostic quality, as these methods lose spatial information and provide limited clinical meaning. Essentially, validating how accurately cast suppression recreates the true underlying anatomy remains fundamentally limited due to the lack of paired cast and no-cast images, precluding direct comparison with ground truth anatomy. An alternative approach for validation is to assess the impact of cast suppression on automated fracture detection performance, potentially demonstrating clinical utility. However, this approach introduces its own methodological concerns. Fracture detection models used for validation may suffer from biases related to their training datasets, especially underrepresentation of radiographs containing cast artefacts. Moreover, the performance metrics of object detection models vary significantly depending on how training and test data are partitioned, with different data splits yielding different performance estimates for the same underlying model architecture [12]. Methods that account for this variability, such as *k*-fold cross-validation, provide more statistically robust estimates of model performance but require training *k* models, making them computationally expensive [13, 14] and often impractical in clinical research settings where resources and time are limited [15–17]. Finally, the nature of the training data used in these studies, such as the imbalance of specific cast types (e.g., non-circumferential splints, fibreglass) also limits generalisability.

Building on prior work in cast artefact suppression and motivated by the limitations of unpaired image translation approaches, we propose a novel method for generating synthetic paired datasets that enable the use of paired image translation methods. Our approach leverages a CycleGAN model to synthesise cast artefacts from real, cast-less radiographs, producing paired images suitable for Pix2Pix training. Using this synthetic dataset, we train Pix2Pix models and objectively compare their performance using spatially preserved metrics. Finally, we assess how synthetic data augmentation and cast suppression models influence fracture detection accuracy, quantifying the relative benefits of synthetic versus real cast data under varying levels of cast image availability.

## Methods

This study was exempt from Human Research Ethics Committee review as a retrospective quality improvement project. It was consistent with the National Health and Medical Research Council (NHMRC) Ethical Considerations in Quality Assurance and Evaluation Activities (2014) guideline in Australia.

### Data curation and hardware

We retrospectively identified patients imaged at a large, tertiary hospital who underwent any wrist radiography between January 1^st^, 2015 and January 20^th^, 2025. This initial search resulted in 31,001 radiographs, which we refer to as the MELBWRI-DX dataset. All wrist radiographs were retrieved from the picture archiving and communication system (PACS) in PNG format and normalised to 8-bit grayscale range (0–255) with zero-padding resizing to 1024 × 1024 pixels. We then separated “cast” and “cast-less” images, where “cast” images refer to follow-up radiographs acquired with casts, and “cast-less” images refer to radiographs without casts.

To ensure a clean dataset of images for synthetic cast generation, we selected a sub-sample of cast-less images using a YOLOv8 classification model. This model was trained exclusively on the annotated and publicly available GRAZPEDWRI-DX dataset [18] for 100 epochs on resized 256×256 images to classify the presence or absence of cast artefacts. The trained model was then applied to our MELBWRI-DX dataset, outputting a probability score (0-1) indicating the likelihood of cast presence for each image. The 10,000 images with the lowest probability of cast presence were retained for subsequent cast suppression modelling to balance computational feasibility with dataset size. The model training details and distribution of classification probabilities can be found in the supplementary information.

The YOLOv8 classification model and all subsequent models in this study were trained using Python (v3.10), Pytorch 2.4, and NVIDIA CUDA 12 libraries, on two NVIDIA Tesla P40 GPUs with 24 GB of VRAM each.

### Synthetic dataset generation and Pix2Pix model training

The generation of cast images was performed using a previously published CycleGAN-based architecture, which included a perceptual loss (PL) function and a self-attention layer (SAL) [10]. This model used a U-Net 512 generator with nine layers each for up- and downsampling, an Adam optimiser with batch size□4, and an initial learning rate α□=□0.002. The model incorporated a perceptual loss computed using a VGG16 network pretrained on ImageNet [19] and a self-attention layer to improve the global coherence of generated images [20]. This pre-trained model was applied to the 10,000 cast-less radiographs from MELBWRI-DX in the reverse direction (generator B→A) compared to its original cast suppression application, producing 10,000 synthetic cast images.

The resulting 10,000 aligned image pairs (cast-less originals paired with synthetic cast versions) were randomly divided into 8,000 training pairs and 2,000 test pairs (80:20 split). Figure 1 shows representative examples of the synthetic training pairs. Three Pix2Pix models were trained using different U-Net generator architectures: U-Net 256, U-Net 512, and U-Net 1024, representing networks with varying architectural complexity and feature extraction capabilities. All models were trained for 100 epochs with a constant learning rate of 0.002 for the first 50 epochs, followed by linear decay over the remaining 50 epochs. The Adam optimiser was used with β□ = 0.5 and batch size of 1.

**Fig. 1.**
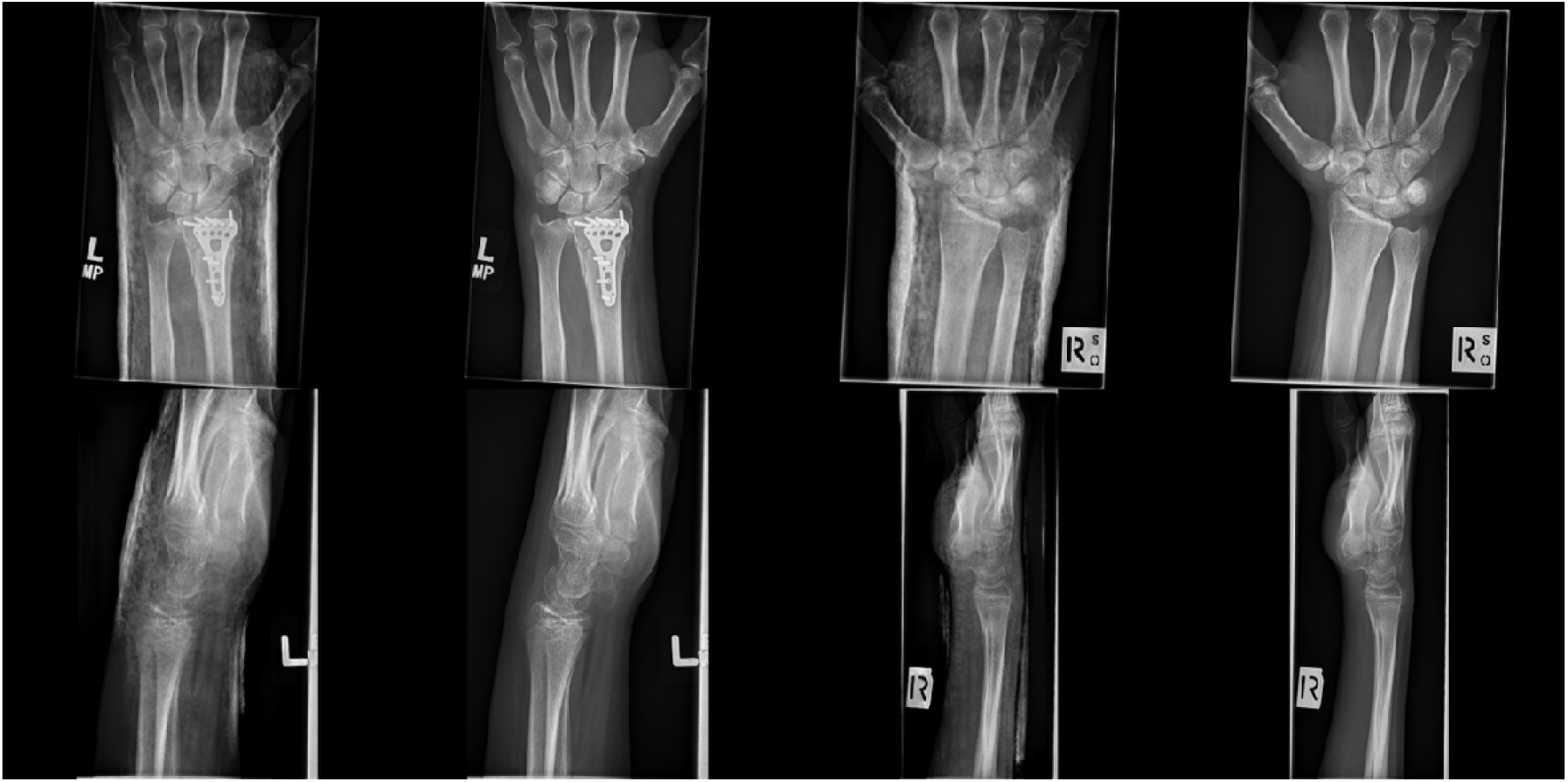
Representative examples of synthetic paired wrist radiographs from the MELBWRI-DX training dataset. Each pair consists of a synthetic cast image generated using the CycleGAN-based architecture (left) and its corresponding original cast-less radiograph (right).

### Quantitative Pix2Pix model evaluation

The three Pix2Pix model configurations were assessed using the held-out test set of 2,000 image pairs. To assess model performance, structural similarity index measure (SSIM), peak signal-to-noise ratio (PSNR), and mean squared error (MSE) were computed against the corresponding ground truth targets. These metrics jointly assess perceptual similarity, signal fidelity, and pixel-wise reconstruction accuracy. For each U-Net generator configuration (256, 512, and 1024), SSIM, PSNR, and MSE were computed across the entire test set, with results reported as median and interquartile ranges (IQR). Although these metrics do not indicate clinical diagnostic quality, they enable objective comparison between architectures and provide a justified basis for selecting the optimal model configuration for subsequent clinical validation.

### Fracture detection model training

The paediatric wrist trauma dataset, GRAZPEDWRI-DX [18], comprising 20,327 annotated images was used to train fracture detection models. The dataset was partitioned using a 70:15:15 stratified split with a random seed (42), yielding 14,207 training images, 3,077 validation images, and 3,043 test images. Cast prevalence was balanced across the splits, with 28.47% in training, 27.46% in validation, and 29.12% in the test set. Patient-level separation was maintained, with 4,263 unique patients in training, 912 in validation, and 916 in the test set, to prevent data leakage.

Six YOLOv11-medium models were trained using identical hyperparameters but with differing training data compositions:

1. Training dataset with 4,045 cast images (“Full”)
2. Training dataset without cast images (“No-cast”)
3. Training dataset with 1,011 cast images (“25% cast”)
4. Training dataset with 2,022 cast images (“50% cast”)
5. Training dataset without cast images, augmented with 1,011 synthetic cast pairs (“25% Aug”)
6. Training dataset without cast images, augmented with 2,022 synthetic cast pairs (“50% Aug”)

All models employed COCO-pretrained weights and conservative data augmentations, including minimal hue variance (0.01), moderate saturation (0.5) and brightness (0.3) adjustments, limited rotation (±5°), translation (0.1), scaling (0.2), and horizontal flipping (0.5). Vertical flipping, mix-up, and copy–paste augmentations were disabled to preserve anatomical fidelity. Training used the Adam optimiser with an initial learning rate of 0.001, a final learning rate factor of 0.01, momentum 0.937, weight decay 0.0005, and cosine learning rate scheduling. The loss function combined classification loss (weight 0.5), bounding box regression loss (weight 7.5), and distribution focal loss (weight 1.5). Models were trained for 100 epochs at 640 × 640 resolution with a batch size of 16, employing a 3-epoch warmup and disabling mosaic augmentation in the final 10 epochs. The 640 × 640 input resolution represents a standard choice in YOLO training that balances computational efficiency with adequate image detail for fracture detection. Details of the test set demographics are shown in Table 1.

**Table 1.** Demographic and clinical characteristics of the test dataset used to evaluate fracture detection models. Values are presented as counts with percentages unless otherwise indicated. Age is reported as median and range.

| Characteristic | n | (%) |
| --- | --- | --- |
| Total images | 3,043 | - |
| Unique patients | 916 | - |
| Gender |  |  |
| Male | 1,781 | 58.5 |
| Female | 1,262 | 41.5 |
| Age (years) | 11 (0-19) | - |
| Laterality |  |  |
| Left | 1,669 | 54.8 |
| Right | 1,374 | 45.2 |
| Cast present |  |  |
| No | 2,157 | 70.9 |
| Yes | 886 | 29.1 |
| Fracture visible |  |  |
| No | 1,059 | 34.8 |
| Yes | 1,984 | 65.2 |

### Training data composition experiment

The first experiment evaluated all six models on the same test set to investigate how training data composition affects fracture detection performance. Models were compared to assess the relative benefits of real cast data versus synthetic augmentation, and to quantify performance differences when cast images are absent, limited, or abundant in training datasets.

### Cast suppression preprocessing experiment

The second experiment assessed how cast suppression preprocessing impacts fracture detection performance across models with different training exposures to real cast images. The corresponding four models (“Full”, “No-cast”, “25% cast”, and “50% cast”) were evaluated under three preprocessing conditions:

1. The unprocessed test dataset (“Original”)
2. The test dataset with unpaired image processing applied to cast images (“CycleGAN”)
3. The test dataset with the U-Net1024 Pix2Pix model applied to cast images (“Pix2Pix”)

For both experiments, model performance was quantified using mean average precision (mAP), a standard object detection metric that measures detection accuracy across different confidence thresholds [21]. We evaluated mAP at intersection over union (IoU) threshold 0.5 (mAP@0.5) and averaged across IoU thresholds 0.5–0.95 (mAP@0.5:0.95) for the fracture class. The IoU threshold determines the minimum overlap required between predicted and ground truth bounding boxes to consider a detection correct. Evaluation used a confidence threshold of 0.001 to retain all potential detections and a non-maximum suppression (NMS) IoU threshold of 0.6 to remove duplicate bounding boxes with substantial overlap. Other annotated classes in the dataset were either too infrequent for reliable statistical analysis (periosteal reaction, bone anomaly, soft tissue, bone lesion, pronator sign) or not clinically relevant to the study objectives (metal, text). Training loss plots for Pix2Pix and YOLOv11 models are provided in the supplementary materials.

### Statistical analysis

For each Pix2Pix U-Net generator configuration (256, 512, and 1024), SSIM, PSNR, and MSE values were calculated for all 2,000 held-out test image pairs and summarised as median (IQR). First, a Shapiro–Wilk test was performed to assess the normality of SSIM, PSNR, and MSE distributions for each Pix2Pix U-Net generator configuration (256, 512, and 1024) and their paired differences. Since each model output is generated from the same set of real images, and the same set of reference images is used for comparison, the resulting data are paired. For non-normally distributed, dependent data with three related groups, a Friedman test is applied to detect global performance differences across models. Where significant effects were observed, Wilcoxon signed-rank tests were used for all three possible model pairings, with p-values adjusted using the Bonferroni method.

For both fracture detection experiments, model performance was evaluated using mAP@0.5 and mAP@0.5:0.95 values. To obtain robust performance estimates and enable statistical comparisons, 20 bootstrap datasets were generated by sampling with replacement from the 3,043-image test set, with each bootstrap sample equal in size to the original test set (N=3,043). All models and preprocessing conditions were evaluated on the same bootstrap samples, producing paired performance data. In the training data composition experiment, global performance differences among the six YOLOv11-medium model configurations were assessed using the Friedman test for non-parametric, dependent data with multiple related groups. Significant effects were explored with Wilcoxon signed-rank tests for all 15 model pairings (6×5/2 combinations), with p-values adjusted using the Bonferroni method to control the family-wise error rate. In the cast suppression preprocessing experiment, within-model comparisons across the three preprocessing conditions (“Original”, “Pix2Pix”, “CycleGAN”) were performed using Wilcoxon signed-rank tests, with p-values Bonferroni-adjusted for the three possible pairwise comparisons per model. All statistical analyses were carried out using Python (v3.12.4).

## Results

### Quantitative Pix2Pix model evaluation

Across the 2,000 held-out test pairs, SSIM, PSNR, and MSE values were calculated for each U-Net generator configuration (256, 512, and 1024) and their distributions are summarised in Table 2 and Figure 2. Figure 3 shows representative examples of cast suppression using the U-Net 1024 model.

**Table 2.**
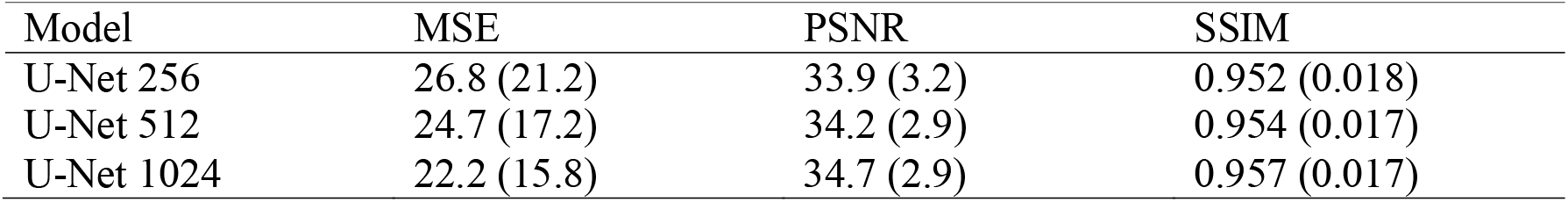
Median (IQR) of SSIM, PSNR, and MSE values across 2,000 held-out test image pairs for each Pix2Pix U-Net generator configuration.

| Model | MSE | PSNR | SSIM |
| --- | --- | --- | --- |
| U-Net 256 | 26.8 (21.2) | 33.9 (3.2) | 0.952 (0.018) |
| U-Net 512 | 24.7 (17.2) | 34.2 (2.9) | 0.954 (0.017) |
| U-Net 1024 | 22.2 (15.8) | 34.7 (2.9) | 0.957 (0.017) |

**Fig. 2.**
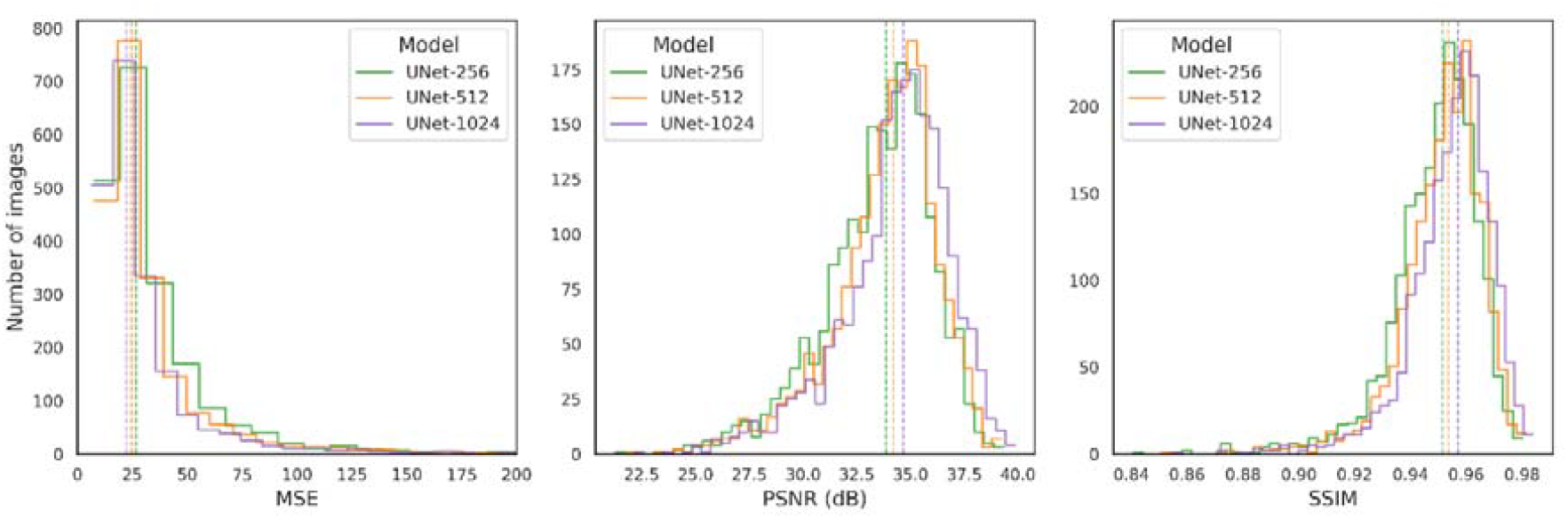
Distribution of MSE, PSNR, and SSIM values for the 2,000 held-out test image pairs, comparing Pix2Pix U-Net generator configurations (U-Net 256, U-Net 512, and U-Net 1024). Vertical dashed lines indicate median values for each configuration

**Fig. 3.**
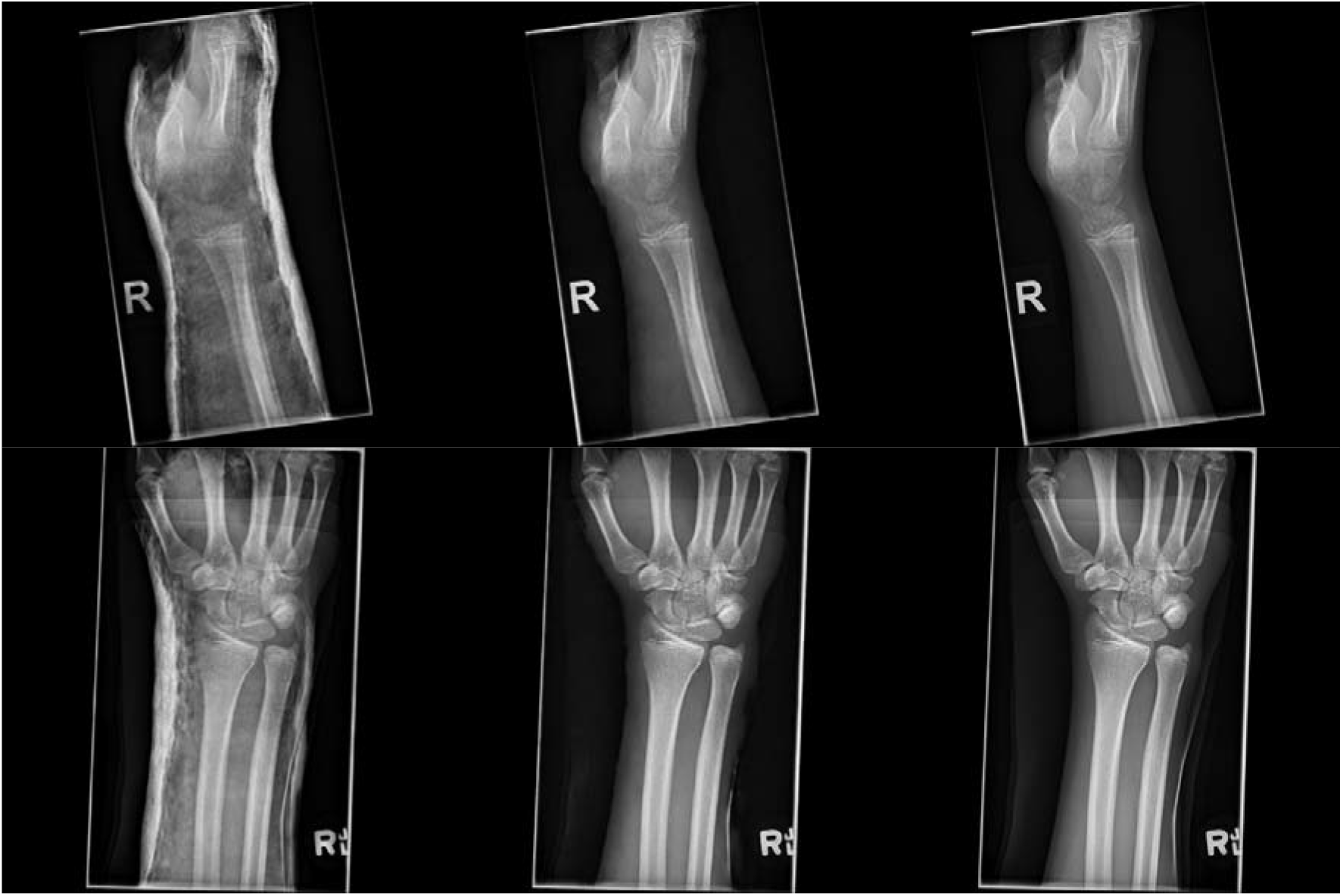
Representative examples of the cast suppression model applied to the MELBWRI-DX test set. The top row shows a lateral projection, and the bottom row shows a posterior-anterior (PA) projection. From left to right: CycleGAN-generated synthetic cast images, U-Net 1024 Pix2Pix cast-suppressed images, and target cast-less reference images.

The U-Net 1024 configuration achieved the highest SSIM values, followed by U-Net 512, with U-Net 256 performing worst. This performance hierarchy was consistent across all metrics, with the U-Net 1024 model producing the highest PSNR and lowest MSE values compared to the smaller configurations. Shapiro–Wilk tests indicated that all metric distributions deviated significantly from normality (p < 0.01), the Friedman test indicated significant effects for all metrics (p < 0.01), and post hoc pairwise Wilcoxon signed-rank tests confirmed significant differences between all model configurations after Bonferroni-adjustment of p-values.

### Training data composition experiment

The training data composition significantly affected fracture detection performance. The “No-cast” model performed worst across both evaluation metrics, while models trained with synthetic cast augmentations (25% and 50%) improved over the no-cast baseline but underperformed compared to models trained with equivalent proportions of real cast images. Among models trained with real cast data, higher proportions yielded better performance, with the “Full” dataset achieving the highest metrics across both thresholds. These performance patterns are illustrated in Figure 4, which shows the distribution of mAP@0.5 and mAP@0.5:0.95 scores across all bootstrap samples for each training configuration. Across the bootstrap samples, global comparisons of the six model configurations showed significant performance differences for both fracture mAP@0.5 (χ^2^(5) = 99.46, p < 0.01) and mAP@0.5:0.95 (χ^2^(5) = 98.91, p < 0.01). Post hoc Wilcoxon signed-rank tests with Holm-adjusted p-values confirmed that all pairwise differences were statistically significant (p < 0.01).

**Fig. 4.**
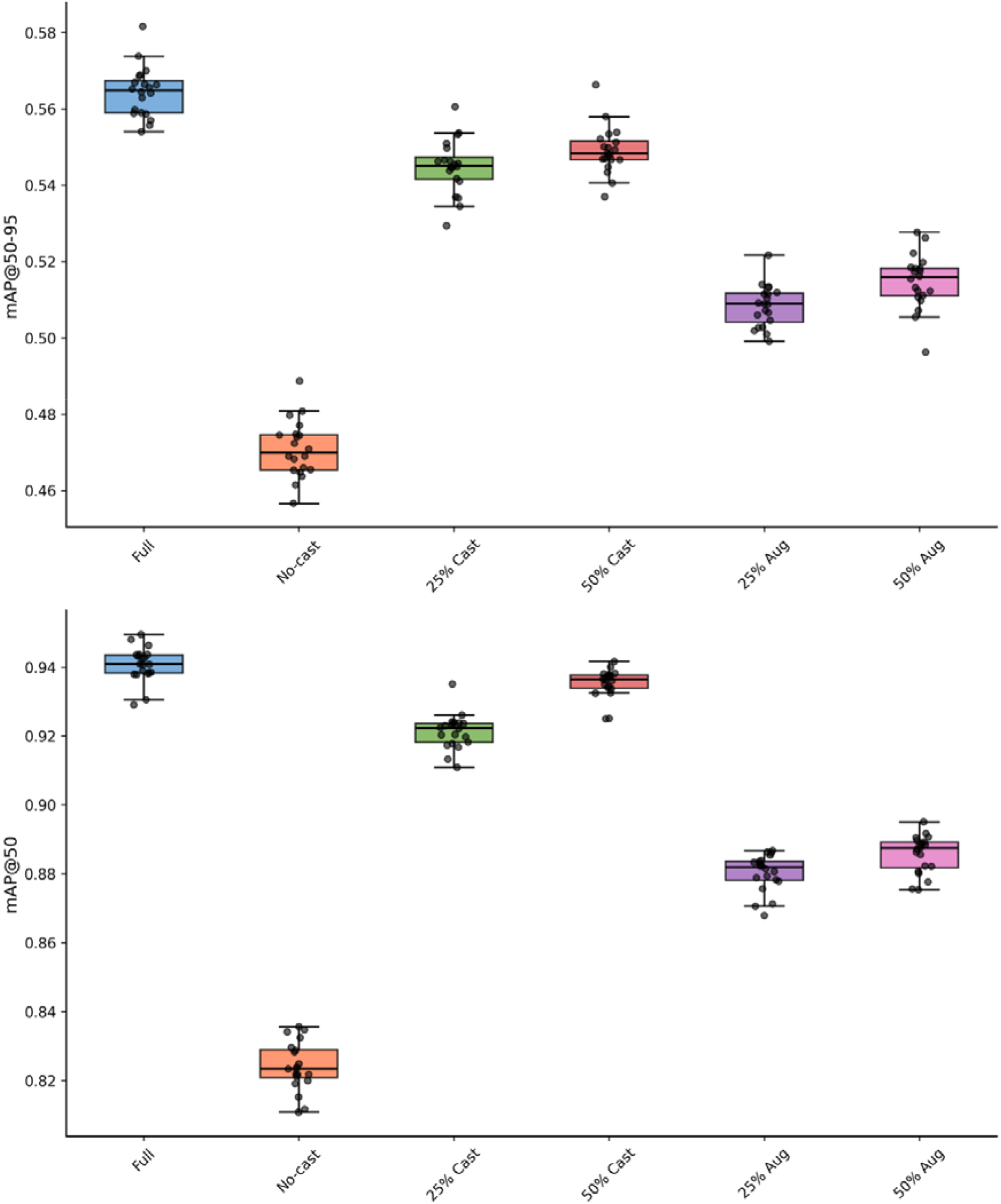
Fracture detection performance comparison across six training data compositions using 20 bootstrap test sets. Box plots show mAP@0.5 and mAP@0.5:0.95 score distributions for models trained with varying proportions of real cast data, synthetic cast augmentation, or no cast exposure

### Cast suppression preprocessing experiment

Cast suppression preprocessing showed conditional effectiveness that depended on model training data composition. For models trained without cast exposure (“No-cast”), both CycleGAN and Pix2Pix preprocessing improved fracture detection performance compared to original images. However, for models already trained with cast data (“Full”, “25% cast”, “50% cast”), cast suppression consistently reduced performance, with original images yielding the highest accuracy. Among suppression methods, Pix2Pix consistently outperformed CycleGAN across all model configurations, as shown in Figure 5. Within-model comparisons using Holm-adjusted Wilcoxon signed-rank tests demonstrated significant differences for most comparisons (p < 0.01). The only exception was the “No-cast” model, where CycleGAN and Pix2Pix showed no significant difference (p = 0.39) in mAP@50, though both outperformed original images.

**Fig. 5.**
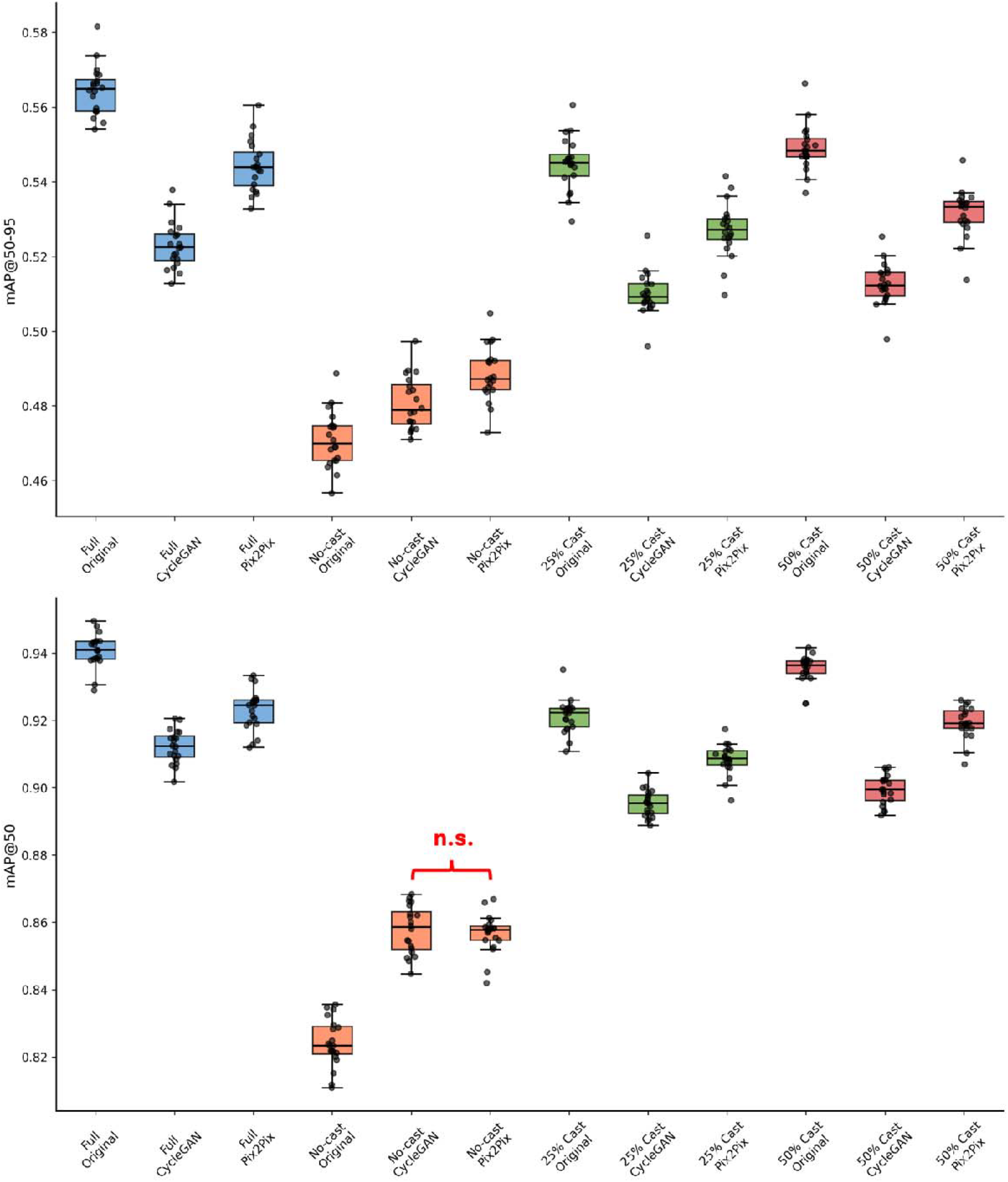
Fracture detection performance of the four YOLOv11 model training configurations (Full, No-cast, 25% cast, 50% cast) across original, CycleGAN, and Pix2Pix test datasets. Box plots show the distribution of mAP@0.5:0.95 and mAP@0.5 scores for each model, evaluated on 20 bootstrap test sets. All within-model pairwise comparisons were statistically significant after Holm adjustment (p < 0.01) except for the “No-cast” model between CycleGAN and Pix2Pix test sets (p = 0.39)

Figure 6 provides visual examples of cast suppression preprocessing, showing CycleGAN and all three Pix2Pix model configurations applied to representative images from the GRAZPEDWRI-DX test set, demonstrating the texture differences between suppression methods and architectures.

**Fig. 6.**
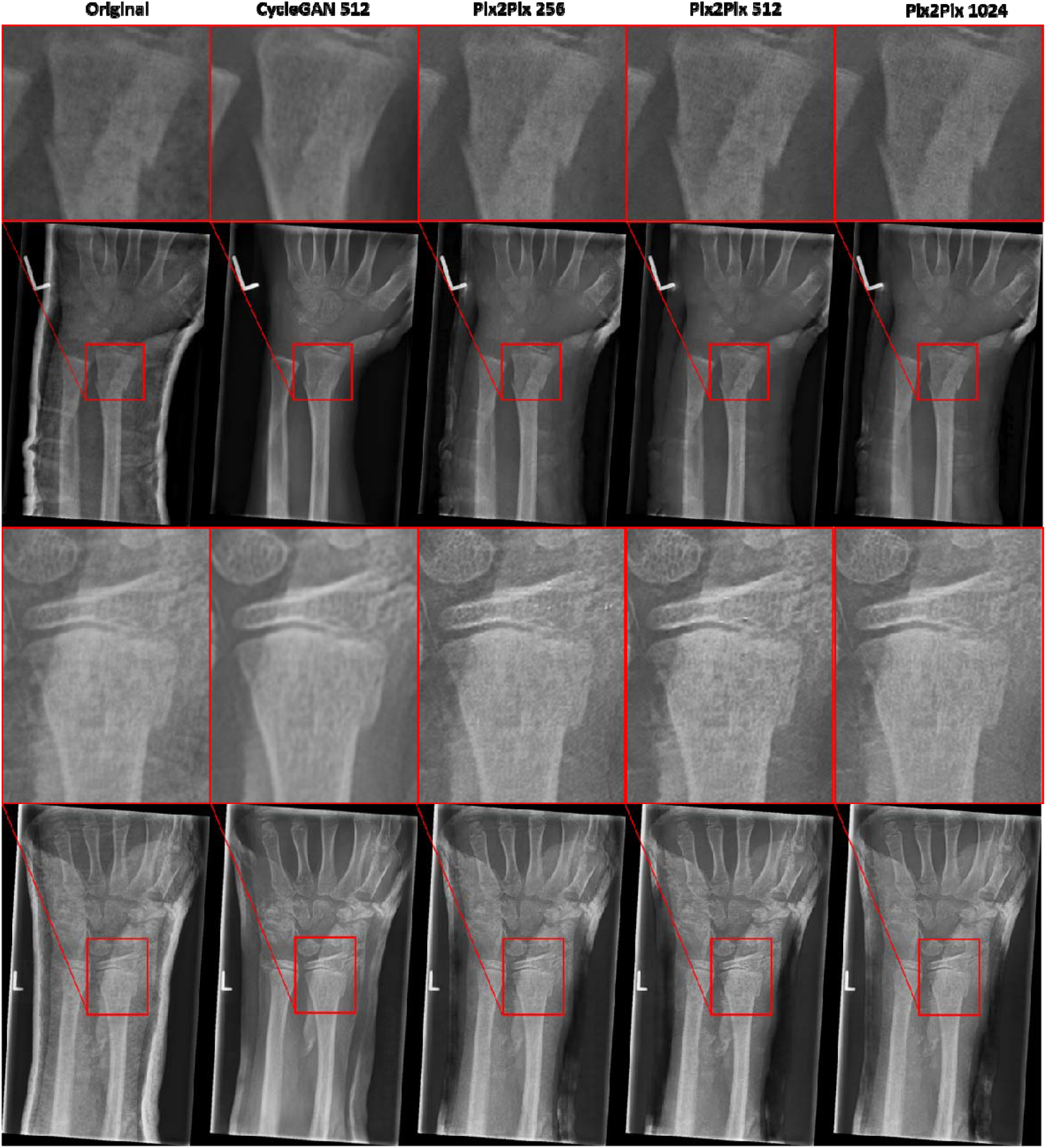
Comparative cast suppression examples across different model architectures applied to GRAZPEDWRI-DX test images. Four rows show two different wrist radiographs, each with the full image (rows 2 and 4) and corresponding fracture detail region (rows 1 and 3). From left to right: original cast image, U-Net 512 CycleGAN cast suppressed image, and cast suppressed images from Pix2Pix models with U-Net 256, U-Net 512, and U-Net 1024 architectures, respectively. Red boxes in full images indicate the location of detailed fracture regions shown in the zoomed panels.

## Discussion

This study addresses key methodological limitations in cast suppression research by generating synthetic paired datasets that enable spatially aware quantitative evaluation. Previous studies have relied on histogram-based similarity measures that lose spatial information [8, 10] or resource-intensive radiologist assessments [9], limiting robust and efficient model comparison. Our paired dataset approach allows direct application of metrics that preserve spatial information, facilitating justified architectural selection. The consistent superiority of the U-Net 1024 Pix2Pix model across all metrics demonstrates the practical value of this quantitative framework for model development.

The training data composition experiment reveals the superior performance of real cast data over synthetic augmentation for fracture detection. While synthetic augmentation improved performance over no-cast baselines, real cast data consistently outperformed synthetic alternatives across all evaluated proportions. The current limitations in generative approaches emphasise the continued need for comprehensive real-world dataset curation. The value of datasets like GRAZPEDWRI-DX extends beyond fracture detection, as they contain annotations for multiple clinical findings including periosteal reaction, pronator sign, and soft tissue abnormalities. However, the low prevalence of these conditions in our test set prevented meaningful analysis of cast suppression effects on their detection, an important area for future investigation given that post-immobilisation monitoring focuses more on healing complications than fracture localisation.

Our preprocessing experiment revealed important nuances in the effectiveness of cast suppression as a preprocessing step. We demonstrated that models trained without cast exposure benefit from suppression preprocessing, while models with real cast exposure during training experience performance degradation when casts are digitally removed. This extends findings from Lee et al., who showed improved fracture detection with cast suppression preprocessing. However, they used a model trained on FracAtlas [22], a general radiograph dataset spanning multiple anatomical regions beyond the wrist and no documented cast information. Our evaluation using GRAZPEDWRI-DX, a paediatric wrist-specific dataset with over 20,000 annotated images, provides more controlled assessment of cast exposure effects and a larger and more specialised dataset for wrist fracture detection. Additionally, our external validation approach, evaluating cast suppression models trained on institutional data (MELBWRI-DX) against GRAZPEDWRI-DX, represents a more challenging validation scenario that better reflects real-world deployment conditions. This conditional effectiveness has significant implications for clinical deployment, as it suggests cast suppression cannot be universally applied without considering the training characteristics of detection models.

Our statistical methodology using bootstrap resampling provides computational advantages over traditional k-fold cross-validation while maintaining robust performance estimates. This approach requires significantly less computational resources, dispensing of the requirement for training k models for k-fold validation, making it more practical for clinical research settings under limited resources.

Several limitations constrain our results. The absence of radiologist assessment prevents measurement of diagnostic image quality, though automated fracture detection provides less subjective evaluation than human assessment and represents a logical first step before a resource-intensive clinical validation. The lack of annotations for our institutional dataset (MELBWRI-DX) prevented evaluation of cast suppression performance on matched data, introducing domain shift effects that may not reflect optimal performance scenarios. Finally, our CycleGAN model was originally developed with a focus on suppression, potentially leaving room for further optimisation of cast generation.

Future research should pursue several complementary directions. Advanced generative approaches including diffusion models or physics-informed simulation using CT-derived radiographs could produce more physically accurate cast artifacts. Anthropomorphic phantom studies could generate real paired datasets for robust validation. Training larger CycleGAN architectures (U-Net 1024) for cast generation is not computationally feasible under limited resources but could improve synthetic dataset quality. Extension to post-immobilisation monitoring applications requires datasets with more annotations for healing complications, displacement, and consolidation status. Cast suppression could particularly benefit non-specialist clinicians in settings that lack specialised paediatric orthopaedic expertise, as described by Lam et al. for treatment decision support [23]. Ultimately, the impact of cast suppression on automated fracture detection models depends on the representation of casts in their training data.

## Conclusion

By generating a synthetic paired dataset, we enabled more robust and meaningful architecture comparisons of cast suppression models and shifted the research challenge from validating unpaired methods to developing diverse and realistic cast generation techniques. In experiments probing the impact of image translation methods on automated fracture detection, we found that CycleGAN-based augmentation and cast suppression preprocessing improved performance only for fracture detection models that had not been exposed to casts during training. Models trained with real cast data consistently outperformed those trained with synthetic alternatives, reinforcing the importance of comprehensive dataset curation for developing clinically robust AI tools.

## Data Availability

All data produced in the present study are available from the authors upon reasonable request and with approval from the Human Research Ethics Committee of Monash Health.

