## Supplementary information for "Enhancing fracture detection in wrist radiographs via paired synthetic data generation"

**Data curation**

A YOLOv8-Large classification model was trained on the publicly available GRAZPEDWRI-DX dataset to distinguish between radiographs with and without cast artifacts. The training dataset comprised 10,000 images (5,000 cast-positive and 5,000 cast-negative) randomly selected and split into training, validation, and test sets using an 80/10/10 ratio. All images were resized to 256×256 pixels without additional data augmentation. The model was trained from scratch (without pre-trained weights) for 100 epochs using a learning rate of 0.002. Following training, the model was applied to classify all 31,001 wrist radiographs, generating probabilistic predictions for cast presence. Images with p_cast_ <0.001 were selected to ensure the curation of a clean x-ray dataset for subsequent cast suppression modelling. The distribution of classification probabilities for the 31,001 radiographs are shown in Figure 1S.

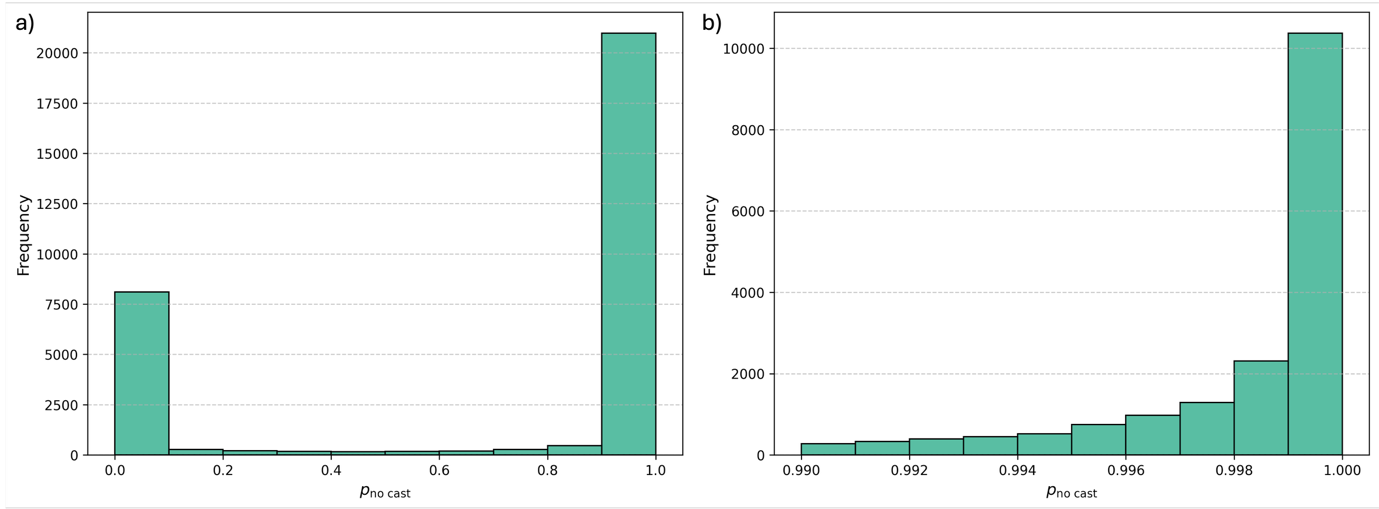

**Fig.1S** Distribution of YOLOv8-assigned “no-cast” probabilities for a) the range 0 to 1, and b) for the range 0.99 to 1. The 10,000 images with the highest probabilities of no-cast were selected for subsequent cast suppression modelling

**Model training**

Pix2Pix models were trained using paired datasets of 10,000 aligned image pairs, each consisting of a real cast-less radiograph and a corresponding synthetic cast image generated by the CycleGAN-based pipeline. The dataset was split into 8,000 training pairs and 2,000 test pairs (80:20 split). Three generator architectures were evaluated (U-Net 256, 512, and 1024), each trained for 100 epochs (50 at a fixed learning rate of 0.002 followed by 50 with linear decay). The Adam optimiser was used with β₁ = 0.5 and a batch size of 1.

Training loss was computed as the sum of the L1 reconstruction loss and adversarial loss, weighted according to the default Pix2Pix configuration. Generator and discriminator losses were logged for all epochs to monitor convergence. Models showed stable training dynamics without mode collapse, with generator loss decreasing steadily towards the end of training and discriminator loss fluctuating within expected bounds for GAN optimisation. Training loss curves for all three Pix2Pix configurations are shown in Figure 2S.

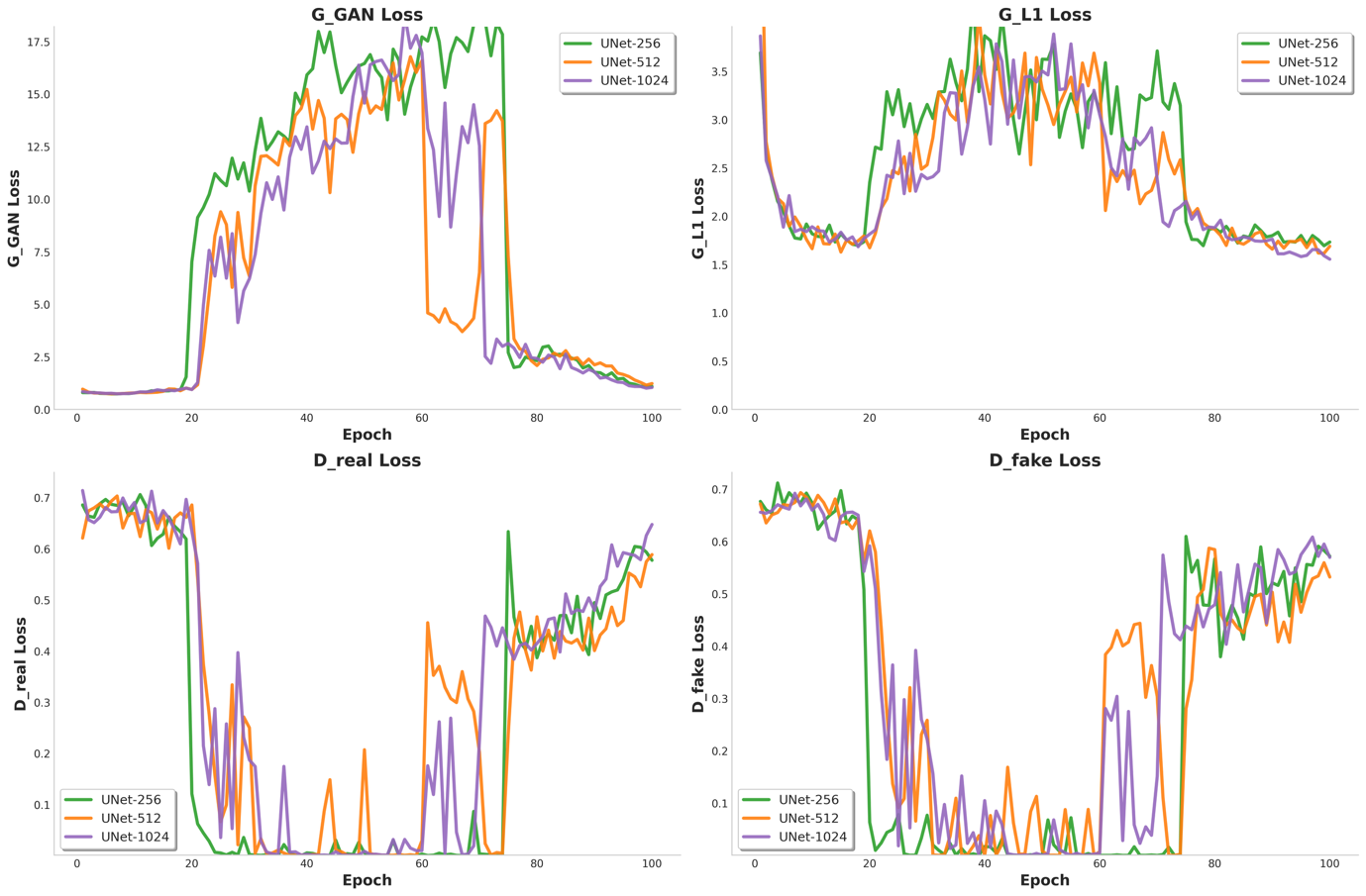

**Fig. 2S** Training loss curves for the three Pix2Pix U-Net generator configurations (256, 512, and 1024). Generator losses are shown with L1 reconstruction and adversarial components, while discriminator loss reflects the adversarial classification task. All models exhibited stable convergence without mode collapse, with generator loss decreasing towards the end of training and discriminator loss fluctuating within expected bounds for GAN optimisation

Fracture detection models were trained using the YOLOv11-medium architecture with COCO-pretrained weights. All six dataset configurations described in the main Methods section were trained using identical hyperparameters, including a batch size of 16, image resolution of 640×640, and 100 training epochs with cosine learning rate scheduling. Losses for classification, bounding box regression, and distribution focal loss were logged for each epoch. Training curves demonstrated consistent convergence across all dataset compositions, with final validation losses stabilising by epoch 80. Loss curves for all fracture detection models are presented in Figure 3S.

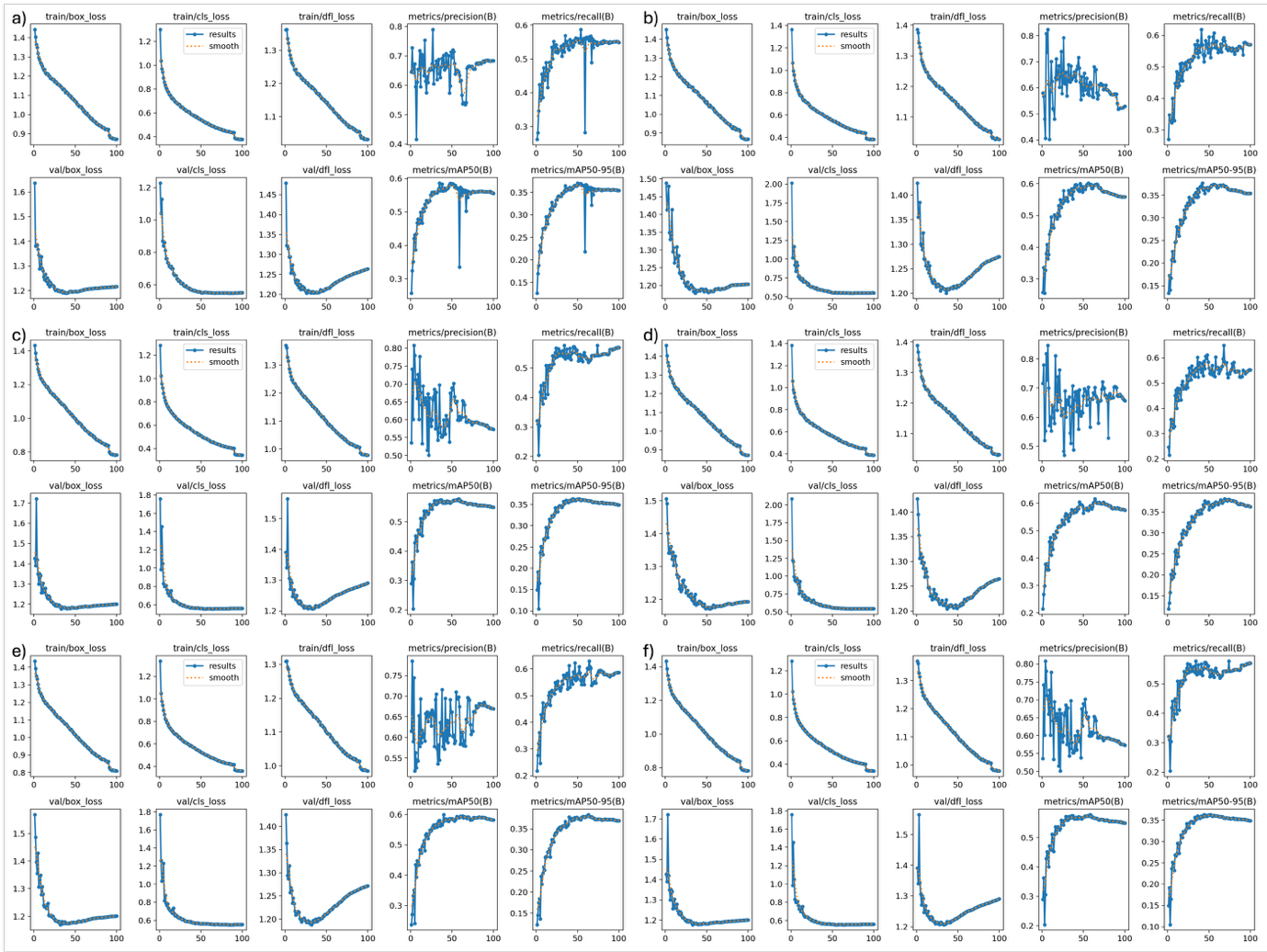

**Fig. 3S** Training and validation metrics for the six YOLOv11-medium fracture detection models: a) Full, b) 25% cast, c) 50% cast, d) No-cast, e) 25% Aug, and f) 50% Aug. Each subplot displays the evolution of bounding box regression loss, classification loss, and distribution focal loss for both training and validation sets, alongside overall dataset precision, recall, mAP@50, and mAP@50:95 (across all 9 classes in GRAZPEDWRI-DX). All models exhibited stable optimisation behaviour, with losses converging and detection metrics plateauing by approximately epoch 80, indicating consistent convergence across dataset compositions
